# A Comprehensive County Level Framework to Identify Factors Affecting Hospital Capacity and Predict Future Hospital Demand

**DOI:** 10.1101/2021.02.19.21252117

**Authors:** Tanmoy Bhowmik, Naveen Eluru

## Abstract

**Background:** As of February 19, 2021, our review yielded a small number of studies that investigated high resolution hospitalization demand data from a public health planning perspective. The earlier studies compiled were conducted early in the pandemic and do not include any analysis of the hospitalization trends in the last 3 months when the US experienced a substantial surge in hospitalization and ICU demand. The earlier studies also focused on COVID 19 transmission influence on COVID 19 hospitalization rates. While this emphasis is understandable, there is evidence to suggest that non COVID hospitalization demand is being displaced due to the hospitalization and ICU surge. Further, with the discovery of multiple mutated variants of COVID 19, it is important to remain vigilant in an effort to control the pandemic. Given these circumstances, the development of a high resolution framework that examines overall hospitalizations and ICU usage rate for COVID and non COVID patients would allow us to build a prediction system that can identify potential vulnerable locations for hospitalization capacity in the nation so that appropriate remedial measures can be planned.

**Method:** The current study recognizes that COVID 19 has affected overall hospitalizations – not only COVID 19 hospitalizations. Drawing from the recently released Department of Health and Human services (DHH) weekly hospitalization data (or the time period August 28^th^, 2020 to January 22^nd^, 2021.), we study the overall hospitalization and ICU usage as two components: COVID 19 hospitalization and ICU per capita rates; and non COVID hospitalization and ICU per capita rates. A mixed linear mixed model is adopted to study the response variables in our study. The estimated models are subsequently employed to generate predictions for county level hospitalization and ICU usage rates in the future under a host of COVID 19 transmission scenarios considering the new variants of COVID 19 and vaccination impacts.

**Findings:** We find a significant association of the virus transmissibility with COVID (positive) and non COVID (negative) hospitalization and ICU usage rates. Several county level factors including demographics, mobility and health indicators are also found to be strongly associated with the overall hospitalization and ICU demand. Among the various scenarios considered, the results indicate a small possibility of a new wave of infections that can substantially overload hospitalization and ICU usage. In the scenario where vaccinations proceed as expected reducing transmission, our results indicate that hospitalizations and ICU usage rates are likely to reduce significantly.

**Interpretation:** The research exercise presents a framework to predict evolving hospitalization and ICU usage trends in response to COVID 19 transmission rates while controlling for other factors. Our work highlights how future hospitalization demand varies by location and time in response to a range of pessimistic and optimistic scenarios. Further, the exercise allows us to identify vulnerable counties and regions under stress with high hospitalization and ICU rates that can be assisted with remedial measures. The model will also allow hospitals to understand evolving displaced non COVID hospital demand.

## BACKGROUND

The Corona Virus Disease (COVID-19) continues to significantly burden social, economic and health systems across the world. As of February 2021, the number of confirmed cases in the world have surpassed 100 million while US alone accounts for about a quarter of these cases^1^. In fact, nearly half of the 27 million cases were reported in the past three months. The total fatalities in the US associated with COVID-19 have crossed 400,000 while daily deaths have averaged nearly 4000 in recent weeks^2^. Further, hospitalization rates have experienced a staggering rise with a peak of about 132 thousand COVID patients hospitalized ^3,4^. Intensive care units (ICUs) are also alarmingly short on space with nearly 33% of the ICUs at a more than 80% occupancy ^5^ and a subset of these ICUs at nearly full occupancy. It is an understatement to suggest that the sustained COVID-19 case numbers and the associated hospitalizations have placed a substantial burden on health care ecosystem comprising of hospitals, clinics, doctors and nurses. The hospital systems have been overwhelmed with COVID-19 cases contributing in the range of 20-50% of the hospitalizations at various facilities. An oft neglected aspect of COVID-19 impacts includes members of the community not contracting COVID-19 but being affected by it. For example, the increasing COVID-19 cases has potentially affected non-COVID hospitalizations i.e. patients are either delaying elective procedures or are being forced to reschedule due to lack of availability in hospitals in some regions. A study conducted in UK ^6^ already anticipated an increase in cancer deaths due to the reduced cancer service triggered by the evolving COVID-19 pandemic. The effect of these delays are likely to extend for months (if not years) after COVID-19 cases are brought under control.

The emergency use authorization of two vaccines (Pfizer-BioNTech and Moderna) offers a potential path to bringing the virus under control. While the initial vaccination rates in the US were slower than anticipated, in recent days, a vaccination rate above 1 million shots per day has been regularly achieved. Even with this impressive vaccination rate, achieving herd immunity could potentially happen only in the early fall months. Further, as new variants of COVID-19 arise with significantly higher transmissibility ^7^, there is a need to maintain our guard in monitoring the cases and resulting consequences (hospitalizations and fatalities). Under these circumstances, there is a need to examine hospitalization and ICU bed usage rates for two reasons. <u>First</u>, to ensure there are adequate hospital facilities for COVID-19 patients until we significantly reduce the threat of the virus. <u>Second</u>, an understanding of hospitalization trends over time will allow us to examine the time when hospital systems can revert to pre-COVID demand. The stabilization of the hospital system will only occur after the displaced health needs of non-COVID patients (delayed surgeries and treatments) are addressed. Towards addressing these aforementioned challenges, the current study develops a comprehensive framework for understanding the critical factors associated with county level hospitalization and ICU usage rates across the US. We estimate the overall hospitalization and ICU usage as two components: (1) COVID-19 hospitalization and ICU per capita rates and (2) non-COVID hospitalization and ICU per capita rates. The consideration of two components (as opposed to the total rate) will allow us to recognize the distinct impact of various factors on each component. The estimated models are employed to generate predictions for hospitalization and ICU usage rates into the future under a host of COVID-19 transmission scenarios considering the new variants of COVID-19 and vaccination impacts.

### Earlier Research And Context Of The Study

A significant amount of research has been conducted on understanding COVID-19 transmission (see^8,9^ for details). However, only a limited amount of research has examined detailed hospitalization data ^10–19^, particularly from a public health planning perspective ^13,14,16,19^. At the planning level, the studies considered the number of people admitted to hospital and ICUs due to COVID-19 as the response variable and employed either time series ^14,16^ or online interactive models ^13,19^ for their analysis. In terms of spatial resolution, different spatial resolutions are explored including region ^14^, county ^19^, country ^13,16^ and city ^13^. With respect to independent variables, the studies found different factors affecting COVID hospitalization and ICU usage rates including demographics (particularly age and racial distribution ^13,19^), mobility trends ^14,19^ and COVID-19 transmission rates ^14,16^. Earlier research efforts on COVID hospitalizations offered several insights. However, given the evolving challenges of COVID-19 the research efforts are still in their infancy.

The current research contributes to the burgeoning literature on COVID-19 hospitalization along multiple directions. <u>First</u>, national level hospitalization data with COVID-19 hospitalization rates at fine spatial resolution have not been easily available. A fine resolution national dataset can allow us to draw insights on the difference across communities that are severely affected relative to other communities. The current research draws on the recently released Department of Health and Human services (DHH) weekly hospitalization data for our analysis. Further, the data employed covers hospitalization data during the peak of the pandemic - from August 28^th^ to January 22^nd^ - with 21 weeks of data across each county.

<u>Second</u>, the current study focuses on understanding the influence of COVID-19 on overall hospitalizations – not only COVID-19 hospitalizations. We recognize that the impact of COVID-19 on overall hospitalizations is going to persist even after the pandemic ends. The displaced hospital demand during the pandemic will need to be addressed once COVID-19 cases reduce substantially. Thus, developing a framework that examines overall hospitalizations and ICU bed usage as a result of COVID and non-COVID hospitalizations can better reflect the plausible hospital system recovery path to pre-COVID level hospitalization trends.

<u>Third</u>, earlier work on hospitalizations employed simple time series models or focused on descriptive and visualization exercises to understand the association between hospitalization and other factors. In our study, we employ a robust modeling framework to analyze the hospitalization and ICU usage demand at a county level. Specifically, we adopt a mixed linear modelling approach ^8^ to account for the repeated observations at the county level (multiple weeks of data). Further, the model estimation exercise is carried out using a comprehensive list of county level independent variables including a) COVID-19 transmission related factors; b) mobility trends; c) health indicators; d) demographics; e) spatial factors and f) temporal factors.

<u>Finally</u>, the model developed is employed to generate predictions (at various spatial units including country, region, state and county) for hospital and ICU usage for COVID and non-COVID patients under a host of future scenarios of COVID-19 transmission. The scenarios are generated considering potential influence of vaccination and the uncertainty associated with COVID-19 variants. The exercise provides an understanding of how hospitalizations and ICU bed usage rates might possibly vary over time across the country.

## EMPIRICAL ANALYSIS

### Data Collection and Preparation

The primary focus of the analysis is to study factors affecting two measures: 1) hospitalization rates (measured as number of beds used) and 2) ICUs used. In our analysis, we examine these measures for COVID and non-COVID patients. Hence, a total of 4 dependent variables are analyzed including: COVID-19 hospitalization and ICU rate per 100K population; and non-COVID hospitalization and ICU rate per 100K population. The data for our analysis is drawn from the DHH database compiled from approximately 5,000 hospitals encompassing 2,462 counties in the country ^20^. The dataset provides information on weekly hospitalization and ICU data from July31^st^, 2020 through January 22^nd^, 2021 (total 32 weeks data). For the analysis, we aggregate the weekly level hospital and ICU data at a county level and consider their natural logarithm to generate the final dependent variables.

In terms of independent variables, six broad categories are considered. An exhaustive list of these variables are presented in Table 1. Within the COVID-19 related factors, we consider different measures including county level weekly COVID-19 transmission rate, percentage change in the virus transmission rate compared to the preceding 3 week average, and if there is a growth in COVID cases in last week. We collect the mobility trends from PlaceIQ ^21^ that provides daily exposure matrices through the smartphone movement data for counties with at least 100 devices reported in a day. For our analysis, we selected the counties with available mobility data resulting in 2,018 counties. Out of these 2,018 counties, there was a small sample of counties (∼250) that do not have any hospitals and are excluded from our analysis. The final dataset consist of total 1,765 counties with 21 weeks of data (from August 28^th^ to January 22^nd^) for each county. The reader would note that these 1,765 counties account for more than 95% of the US population.

**Table 1.** Descriptive Statistics of the Dependent and Independent Variables.

| Variables | Source | Mean | Min/Max | Sample Size |
| --- | --- | --- | --- | --- |
| **Dependent Variables** |  |  |  |  |
| Ln (COVID hospitalization rater per 100k) | DPH<sup>a</sup> | 1.935 | 0.000/7.525 | 37,065 |
| Ln (non COVID hospitalization rater per 100k) | DPH | 4.069 | 0.000/8.341 | 37,065 |
| Ln (COVID ICU rater per 100k) | DPH | 0.752 | 0.000/6.995 | 37,065 |
| Ln (non-COVID ICU rater per 100k) | DPH | 1.536 | 0.000/6.752 | 37,065 |
| **Independent Variables** |  |  |  |  |
| ***COVID-19 related factors*** |  |  |  |  |
| COVID case per 100 people, 1 week lag | CSSE | 5.008 | 0.000/4.560 | 37,065 |
| COVID case per 100 people, 2 weeks lag | CSSE | 5.008 | 0.000/4.560 | 37,065 |
| Difference from 3 week moving average | CSSE<sup>b</sup> | 0.086 | -1.000/2.002 | 37,065 |
| Weekly COVID-19 cases higher than the moving average | CSSE | 0.587 | 0.000/1.000 | 37,065 |
| ***Mobility Trends*** |  |  |  |  |
| Ln (Daily Average Exposure), 2 weeks lag | CEI<sup>c</sup> | 4.536 | 2.319/6.841 | 37,065 |
| Ln (Daily Average Exposure), 3 weeks lag | CEI | 4.521 | 2.324/6.841 | 37,065 |
| ***Demographic Characteristics*** |  |  |  |  |
| Young people percentage | ACS<sup>d</sup> | 22.403 | 7.155/35.987 | 1,765 |
| Hispanic percentage | ACS | 10.015 | 0.653/96.322 | 1,765 |
| African American percentage | ACS | 9.720 | 0.113/76.331 | 1,765 |
| Female percentage | ACS | 50.348 | 37.041/56.145 | 1,765 |
| Ln (median income) | ACS | 10.866 | 10.149/11.821 | 1,765 |
| Income inequality ratio (80<sup>th</sup> /20<sup>th</sup> percentile) | CHRR<sup>e</sup> | 4.540 | 2.987/9.148 | 1,765 |
| ***Health Indicators*** |  |  |  |  |
| Asthma % for >= 18 years | CDC | 9.417 | 7.400/12.300 | 1,765 |
| Ln (number of cardiovascular patients per 1000 Medicare beneficiaries) | CHRR | 4.119 | 3.157/4.891 | 1,765 |
| Hepatitis C Cases per 100K people | CDC<sup>f</sup> | 1.064 | 0.000/5.600 | 1,765 |
| Ln (HIV rate per 100K People) | CDC | 4.780 | 0.723/7.859 | 1,765 |
| Ln (cancer rate per 100K People) | CDC | 6.119 | 5.489/6.436 | 1,765 |
| **Spatial factors** |  |  |  |  |
| West region | USA map | 0.120 | 0.000/1.000 | 1,765 |
| Mid-West region | USA map | 0.108 | 0.000/1.000 | 1,765 |
| North-East region | USA map | 0.308 | 0.000/1.000 | 1,765 |
| Top 10 tourist state | CHRR | 0.252 | 0.000/1.000 | 1,765 |
| Number of airports per 100k people | CHRR | 1.269 | 0.000/24.927 | 1,765 |

<sup>b</sup> = Department of Health and Human services <sup>20</sup>; <sup>b</sup> = Center for Systems Science and Engineering Coronavirus Resource Center at Johns Hopkins University <sup>22</sup>; <sup>c</sup> = COVID Exposure Indices <sup>21</sup>; <sup>d</sup> = American Community Survey; <sup>e</sup> = County Health Rankings & Roadmaps; <sup>f</sup> = Central for Disease Control System.

A visual representation of salient characteristics of the data are presented in Figure 1. Specifically, for US and West region, we present a) hospitalization and ICU rate (total and by COVID patients), b) COVID-19 transmission rates, and c) average mobility exposure (Other regions’ summary are in the supplemental material, Figure A.1). From the figure, we can observe how hospitals are inundated with COVID patients while the number of non-COVID patients are declining across the country. The situation is worse in the West region as hospital beds and ICU units are occupied with the influx of COVID-19 patients. In particular, the ICU capacity in the west region dropped to 10% with an availability of only 3 ICU beds per 100k people. All of these statistics clearly depict the challenging situation impacting the hospital sector, even after 1 year into the pandemic.

**Figure 1:**
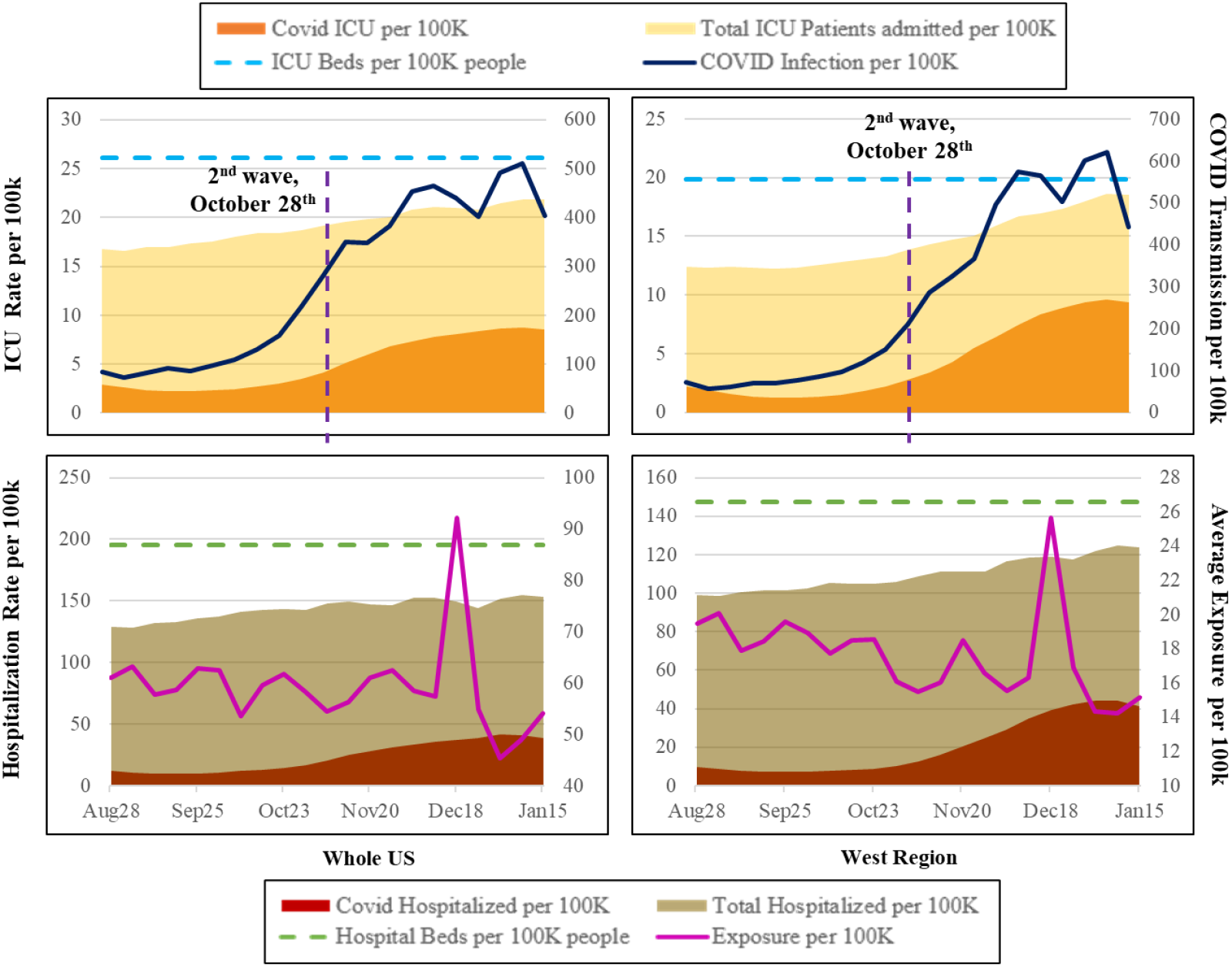
A Representation of the Hospitalization Trends Across the Country and West Region

### Analysis Method

All the response variables considered in the study are continuous in nature and thus, linear regression framework would be an appropriate choice for analyzing the data. However, in our data, every county is considered 21 times and a simple linear regression method is not appropriate for such repeated measures^8^. Therefore we adopted a linear mixed approach (Autoregressive moving average -ARMA model structure, see ^8^ for details) to accommodate for the influence of repeated observations at a county.

## RESULTS

Table 2 provides the COVID and non-COVID hospitalization model estimates. We restrict ourselves to a discussion of COVID hospitalization models. The discussion of the results for the non-COVID hospitalization rate (Section A.1), and estimates for ICU rate models (Table A.1) are included in the supplemental materials.

**Table 2:** Hospitalization Model Results

|  | **COVID Hospitalization** |  | **Non COVID Hospitalization** |  |
| --- | --- | --- | --- | --- |
| Parameter | Estimate | t-statistics | Estimate | t-statistics |
| Intercept | -20.151 | -11.971 | -9.858 | -6.025 |
| ***Covid-19 Transmission Factors*** |  |  |  |  |
| COVID case per 100 people, 1 week lag | --** | -- | -0.076 | -4.292 |
| COVID case per 100 people, 2 weeks lag | 1.266 | 17.790 | -0.107 | -5.933 |
| x Effect in the West Region | -0.145 | -1.765 | -- | -- |
| x Effect in the South Region | -0.694 | -8.575 | -- | -- |
| Weekly COVID-19 cases higher than the moving average (base is covid-19 cases same or lower) | 0.058 | 5.753 | -- | -- |
| % difference with the preceding 3 week moving average | 0.089 | 3.986 | -- | -- |
| x Effect in the Mid-West Region* | 0.177 | 5.894 | -- | -- |
| x Effect in the South Region | -0.038 | -1.653 | -- | -- |
| ***Mobility Trends*** |  |  |  |  |
| Ln (Daily Average Exposure) with 2 weeks lag | 0.157 | 7.285 | -- | -- |
| Ln (Daily Average Exposure) with 3 weeks lag | 0.295 | 12.502 | -- | -- |
| ***Demographics*** |  |  |  |  |
| Young population percentage (19 years or less) | -0.051 | -5.511 | -0.043 | -4.835 |
| Hispanic percentage | 0.025 | 11.141 | 0.008 | 3.426 |
| African American percentage | 0.020 | 9.431 | 0.002 | 1.844 |
| Female percentage | 0.173 | 10.719 | 0.153 | 9.758 |
| Ln (median income) | 0.619 | 5.250 | -- | -- |
| ***Health Indicators*** |  |  |  |  |
| Ln (number of cardiovascular patients per 1000 Medicare beneficiaries) | 1.133 | 9.215 | -- | -- |
| Hepatitis C Cases per 100K people | 0.070 | 2.624 | -- | -- |
| Ln (HIV rate per 100K People) | -- | -- | 0.236 | 6.307 |
| Ln (cancer rate per 100K People) | -- | -- | 0.973 | 3.747 |
| ***Spatial Factors*** |  |  |  |  |
| Region (Base: West, South, Pacific) |  |  |  |  |
| Mid-West region |  |  | 0.111 | 1.897 |
| North East region | -0.109 | -2.197 | -- | -- |
| x Effect Since 2nd Wave started (October 30<sup>th</sup>) | 0.204 | 3.923 | -- | -- |
| Top10 tourist state | 0.243 | 3.736 | -- | -- |
| x Effect Since 2nd Wave started | -0.114 | -3.061 | -- | -- |
| Number of airports per 100k people | 0.017 | 1.651 | -- | -- |
| ***Temporal Factors*** |  |  |  |  |
| Effect Since 2nd Wave started (October 30<sup>th</sup>) | 0.360 | 18.212 | -- | -- |
| Effect Since 25<sup>th</sup> December | 0.106 | 6.422 | -0.041 | -3.786 |
| ***Correlations*** |  |  |  |  |
| $\sigma^2$ | 1.912 | 49.925 | 1.370 | 40.445 |
| $\rho$ | 0.930 | 453.710 | 0.965 | 791.200 |
| $\Phi$ | 0.835 | 237.303 | 0.888 | 306.605 |

\*The indented variable starting with “x” is adopted to indicate that the variable represents the interaction term with that specific variable. \*\* the variable is insignificant at 90% significance level.

### COVID-19 Hospitalization Model

#### COVID-19 Related Factors

The consideration of COVID-19 transmission rates in the model recognizes the delay of about 5 to 14 days between transmission and hospitalization. Hence, in our analysis we tested COVID-19 transmission variable with lag of 1, 2 and 3 weeks. Of these lag variables, the 2 weeks lag offered the best model fit. As expected, increase in COVID-19 transmissions is associated with increased hospitalization rate. Further, we find that the impact of transmission rates is lower (yet positive) in the West and South region relative to the rest of the country. In addition, in our analysis, to represent rising COVID-19 cases in the county, we defined several variables such as a) an indicator variable for increasing cases defined as number of weekly cases greater than the 3 week moving average (3WMA) and b) a percentage difference variable representing change in weekly cases relative to the 3WMA. These variable impacts follow expected trends. The indicator variable for increasing cases contributes to increasing hospitalization rates. The difference variable (can be positive or negative) indicates that hospitalizations are sensitive to percentage changes in COVID-19 transmission. The sensitivity is substantially higher for Mid-West region while being slightly lower for the South region.

#### Mobility Trends

Consistent with earlier research, our analysis also highlights how increased mobility in the county results in higher hospitalization rates. We recognize that the impact of mobility on hospitalizations has a lagged effect. Hence, we tested weekly mobility with 2 weeks and 3 weeks lag. The two lag variables offer significant and intuitive results highlighting the role of mobility in addition to the impact of COVID-19 cases.

#### Demographics

With respect to demographic characteristics, we find that counties with higher percentage of young population (aged 19 years or less) are likely to have lower hospitalization rates ^23^. The results for ethnicity composition variables also offer expected results. Counties with higher Hispanic and African American populations are likely to have higher hospitalizations, perhaps attributed to their residence in densely populated neighborhoods and pre-existing chronic medical conditions ^23^. Interestingly, in our analysis, hospitalization rates are found to be higher for counties with higher female population. Finally, median county income is also positively associated with hospitalization rates, possibly manifesting the impact of access to hospitals in these counties.

#### Health Indicators

Consistent with earlier findings, our analysis also shows a significant positive association between the COVID hospitalization rate and pre-existing health risk factors, especially for population with higher incidence of cardiovascular disease and hepatitis C ^24^.

#### Spatial Factors

In our analysis, we tested several spatial factors to account for inherent regional differences in hospital infrastructure, transportation infrastructure, tourism activity, weather patterns and regional culture. The results shows that north east region is likely to experience lower hospitalization rate due to COVID-19 compared to the rest of the country prior to October 30^th^. However, after October 30^th^, the north east region experienced slightly higher hospitalization rates. The higher hospitalization rate for this time period is closely aligned with the increasing caseloads across the country. Further, we considered the tourism status of the state in our analysis by identifying the top and bottom 10 desirable states with respect to tourism activity. As expected, we find a positive effect of the top 10 tourist attraction state on the COVID hospitalization rate, perhaps indicative of the higher virus transfer in these regions. However, the effect was marginally reduced from October 30^th^. Finally, the variable specific to the number of airports per 100k people reveals a positive impact on the COVID hospitalization rate.

#### Temporal Factors

With respect to temporal variables, we consider various functional forms including continuous (linear, square and other polynomial forms of week difference) and indicator variables (such as Pandemic effect since October 30^th^ (second wave), since mid of December and from 25^th^ December and later). As expected, we find a positive effect of the second wave variable indicating a higher hospitalization rate across the country from 30^th^. The results also indicate that hospitalization rate for COVID patients accelerated further from 25^th^ December.

#### Correlation Factors

The significant results for the parameters *σ*^2^, *ρ* and *ϕ* clearly highlight the presence of common unobserved factors affecting the county COVID-19 hospitalization rate across multiple observations.

## DISCUSSION

### Hospital demand projection^1^

The main objective of the current study is to contribute to public health planning by evaluating how hospitalizations trends evolve over time. Specifically, the focus is on drawing insights on future demand while accommodating for the inherent uncertainty in future COVID-19 transmission. In terms of future COVID-19 transmissions, the reader would note that while

COVID-19 virus transmissions are receding currently, the emergence of highly transmissible variants might affect the trajectory. Towards this end, we consider four potential COVID-19 evolution scenarios as follows ^25^:

1. *<u>Peak and Valley (PV)</u>*: In this scenario, COVID-19 cases are assumed to follow a series of repetitive COVID-19 waves (ups and downs) throughout the spring of 2021 and beyond. The waves could be a result of delays in vaccination production and/or vaccination drives resulting in a longer timeframe for COVID-19 cases to reduce.
2. *<u>Unexpected Third Spike (UP)</u>*: In the presence of new variants of COVID-19, a potential spike in COVID-19 transmissions that might occur in March or April is considered in this scenario. In this scenario, while the spike results in increased transmission, active vaccination drive can contribute to case reduction starting from the end of April.
3. *<u>Slow Burn (SB)</u>*: In this scenario, we assume that the cases will go down slowly in the coming months while not diminishing completely until the end of July.
4. *<u>Rapid Vaccination (RV)</u>*: It is possible that multiple vaccine candidates might be approved (such as Johnson and Johnson and others) in the near future rapidly increasing US vaccination rates. In this scenario, COVID-19 transmission rates are likely to reduce and reach a very manageable level early summer.

The first two scenarios represent a pessimistic outlook towards COVID-19 transmission while the scenarios 3 and 4 represent an optimistic outlook. Figure 2 provides a representation of future COVID-19 transmission for the four scenarios described. The scenarios are generated employing percentage changes in cases at a county level thus ensuring spatial variability in the country. The spectrum of scenarios will enable us to examine the variation in hospital bed and ICU demand in the future. The reader would note here that mobility trends will also influence COVID-19 hospitalization demand. However, to focus on COVID-19 transmission trends, we consider the same mobility profile across all four scenarios (Figure A.2).

**Figure 2:**
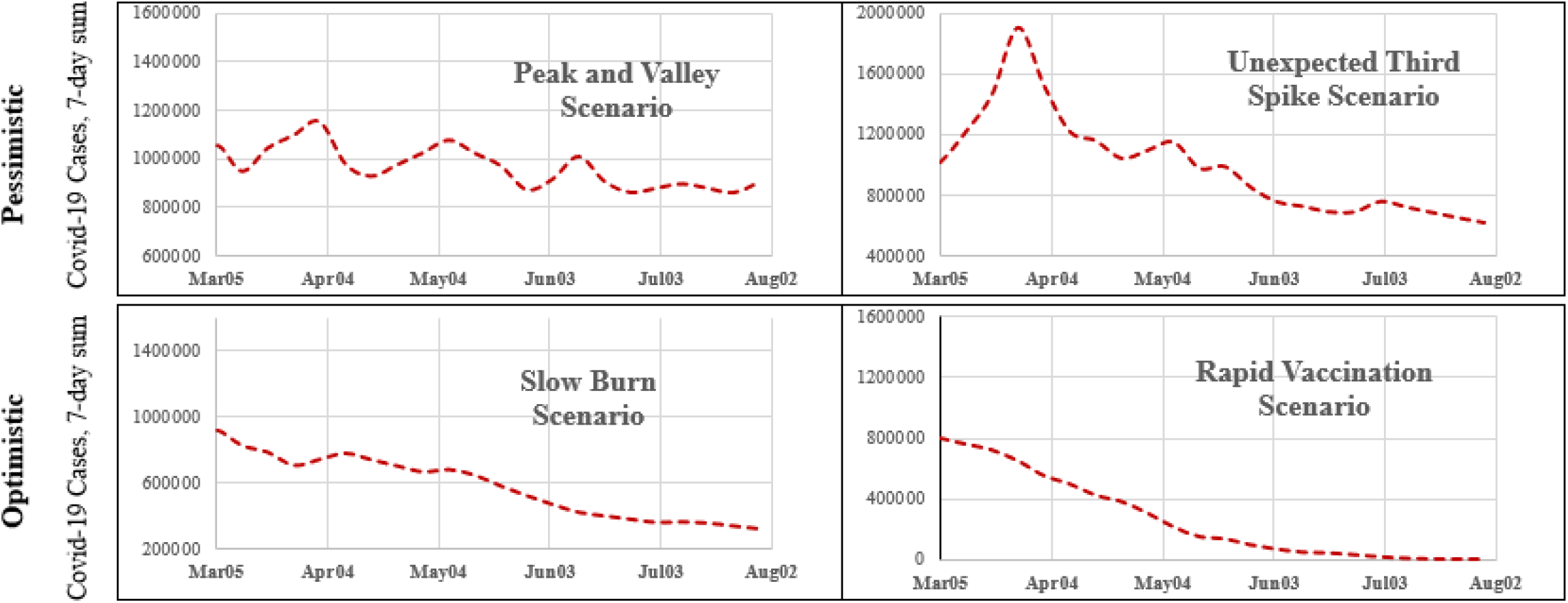
A Representation of the Assumed Scenarios of the COVID-19 Transmission Rate in Future

For these four scenarios, we forecasted hospital and ICU capacity using the proposed linear mixed model for all counties considered in the analysis over the next 22 weeks (March 5^th^ to August 6^th^). The model predictions at the county level can be appropriately aggregated to any spatial resolution to provide insights at the national, regional, state and county level.

A national and regional outlook for hospitalization and ICU rates are provided in Figure 3a and 3b for the scenarios. In this set of figures, we present results for US, North-East and West regions for ease of presentation (results for other regions are presented in supplemental material – Figures A.3, and A.4). The results for all the scenarios follow expected trends with the pessimistic scenarios showing higher demand and optimistic scenarios presenting with lower demand. The national results also clearly indicate that national hospital supply can meet the demand under all scenarios. However, it is important to recognize that excess demand in a county/region cannot be transferred to a county/region with deficit. An examination of the hospitalization and ICU rates in North-East and West reinforces this point. Under pessimistic scenarios, capacity in the North-East and West regions might come near the maximum available capacity (≥ 90%). Specifically, North-East region might experience a supply-demand mismatch for hospital beds while ICU supply-demand mismatch is expected to more likely in the West region. These results reflect inherent regional and demographic differences across the country.

**Figure 3a:**
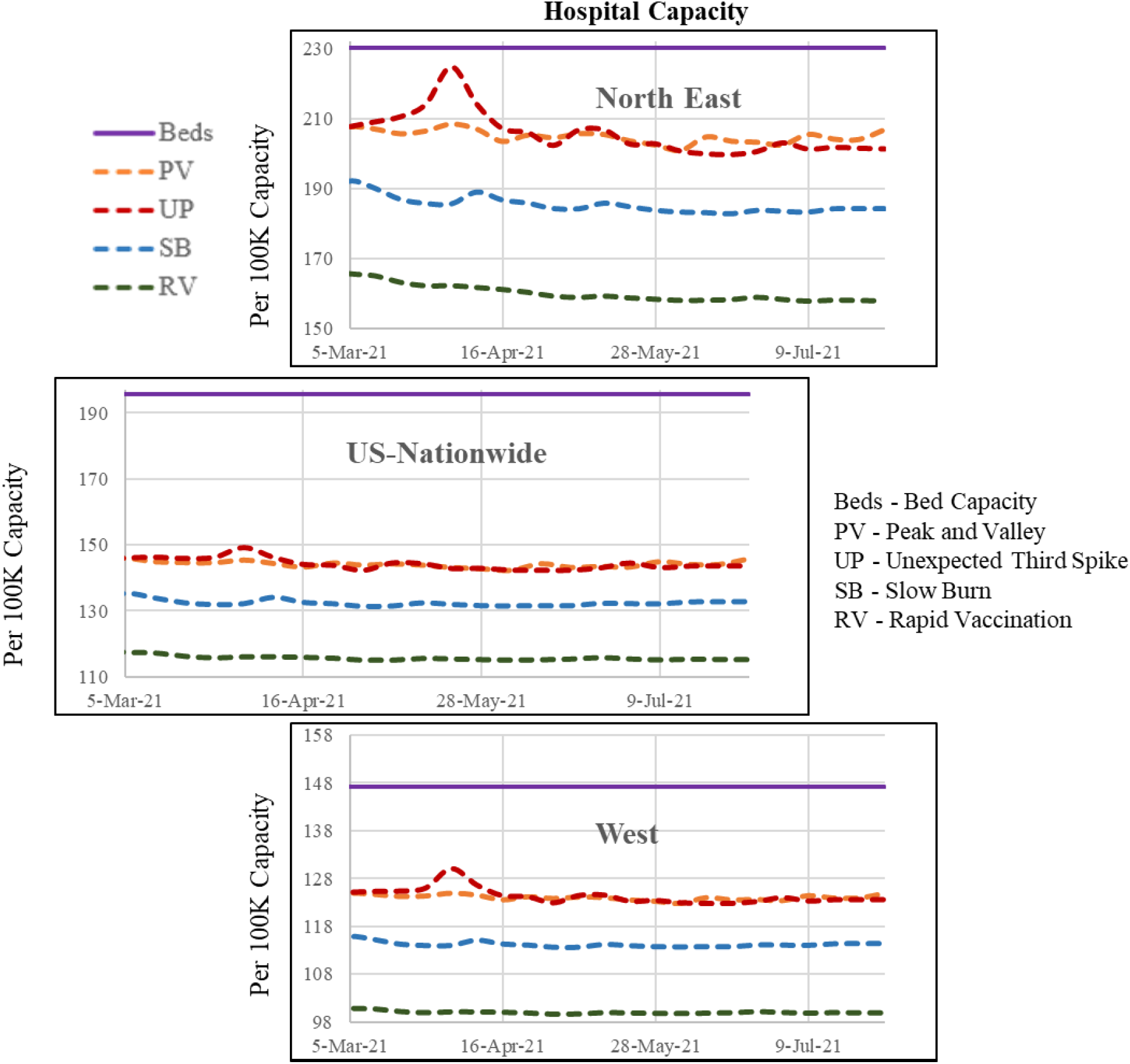
Future Hospital Capacity Across the Country and Regions (West and North-east) Based on the Hypothetical Scenarios

**Figure 3b:**
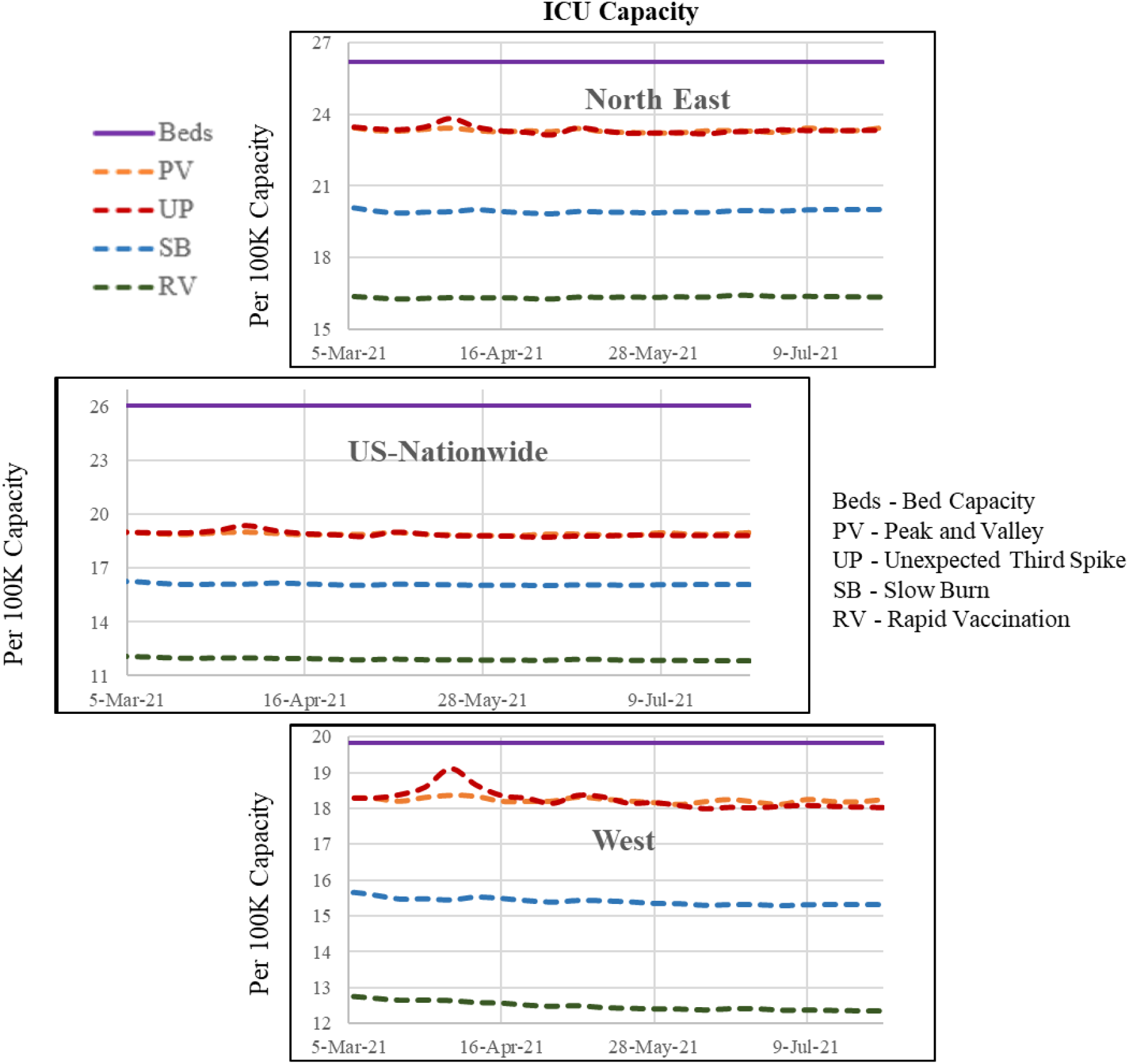
Future ICU Capacity Across the Country and Regions (West and North-east) Based on the Hypothetical Scenarios

To further understand the potential mismatch in demand and supply, we examine what percentage of the counties (out of 1,765) can potentially experience a supply and demand mismatch – defined as at least 90% hospital beds and ICU units being used. We also identify the percentage of counties with at least 25% of hospital beds or ICU units allocated to COVID-19 patients for the four scenarios. The results of this exercise, presented in Figure 4, indicate that hospitalization rates exceed 90% in a range between 13-30% while for ICU usage, the rate vary from 3.5-5.5% of counties across the four scenarios. The number of counties with more than 25% COVID-19 hospitalizations (3-16%) or ICU usage (27-50%) varies significantly across the scenarios.

**Figure 4:**
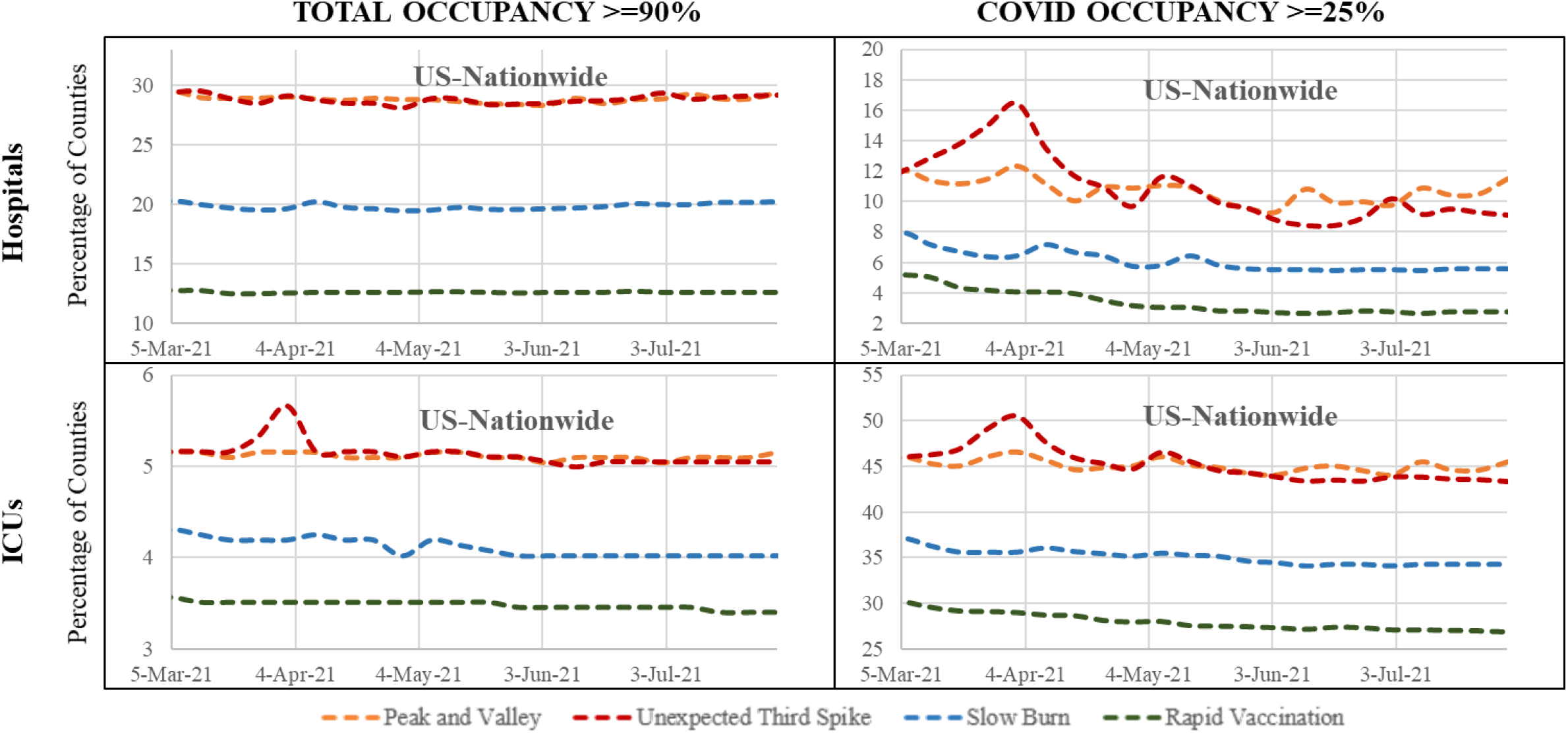
Number of Counties with Capacity over 90% and COVID Patients Over 25%

The results presented so far examined the trends over time. To illustrate our model applicability for county level analysis at a specific time point, we present the results of two scenarios - PV and RV – in July across the states of California and Florida in Figures 5a and 5b. The figures provide the following measures: a) county level hospitalization and ICU capacity usage, and b) number of counties within each category. As expected, higher number of counties are at risk of exceeding capacity in the PV scenario compared to the RV scenario in both states. The figures show how employing the proposed model system, counties at risk at any future time point can be identified to suggest remedial measures such as increasing the staff resources and/or hospital resources as needed. A state by state analysis of hospital capacity usage is presented in Figure A.5.

**Figure 5a:**
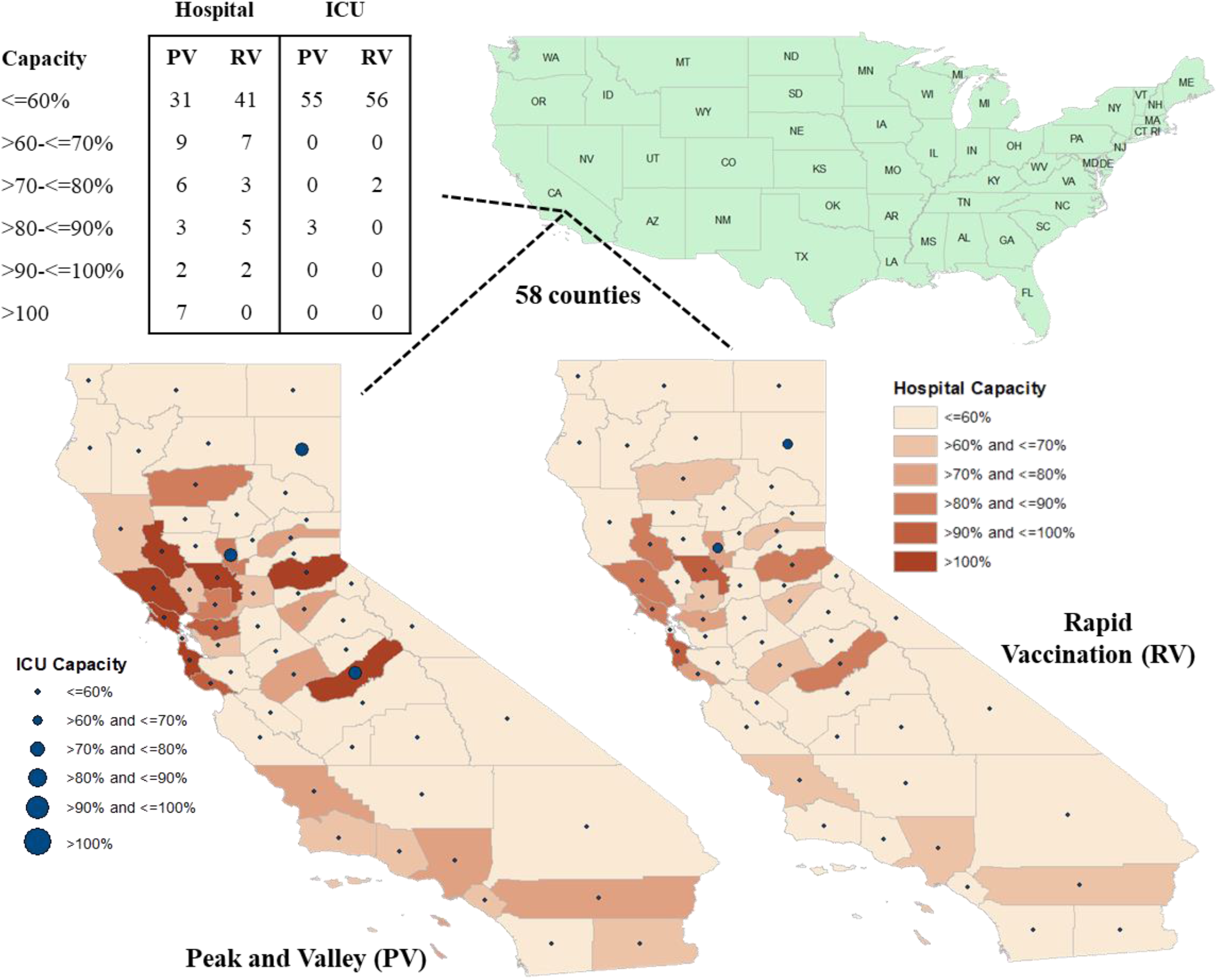
Future Hospital Capacity at Counties (California) Based on the Hypothetical Scenarios

**Figure 5b:**
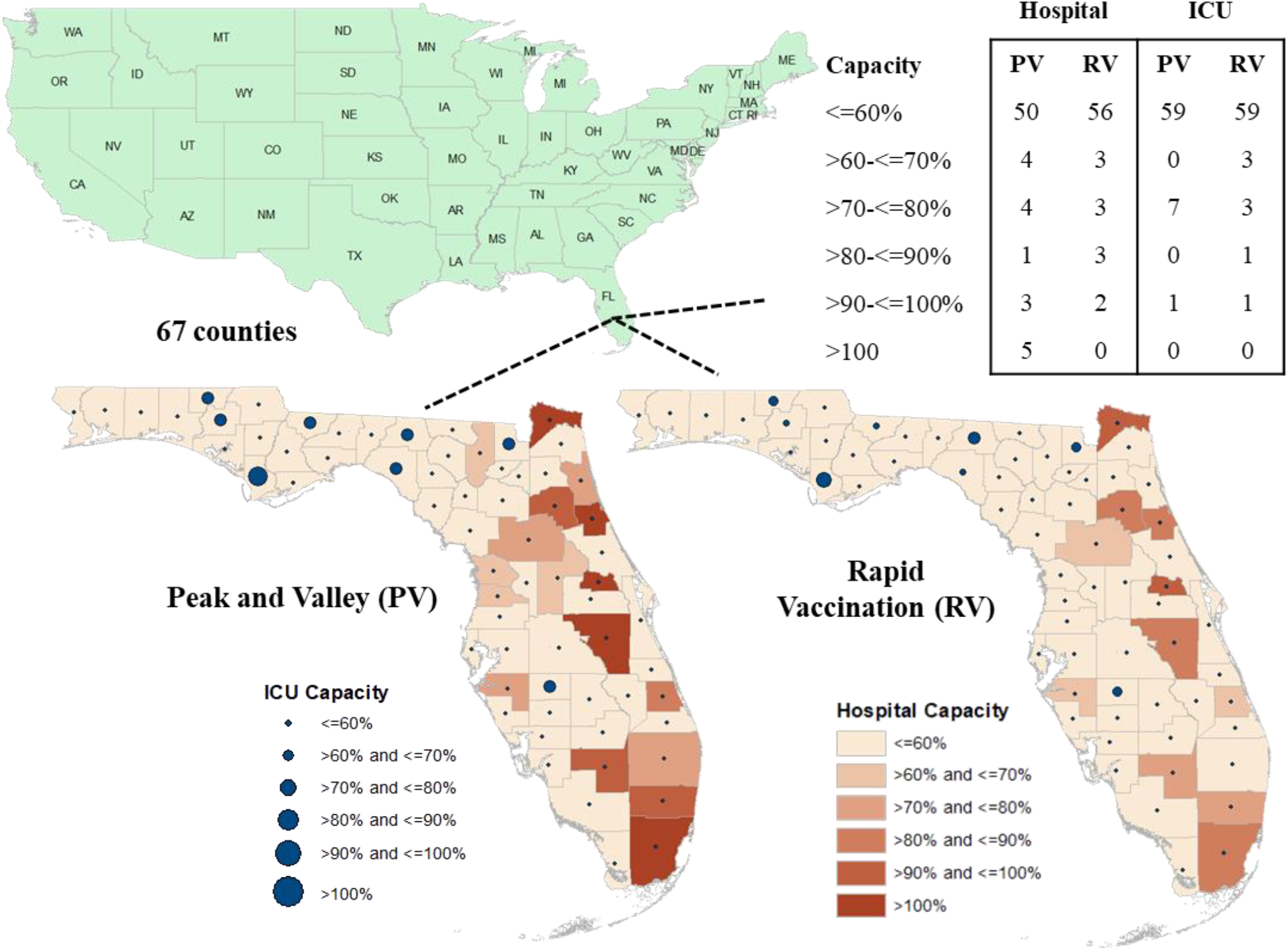
Future Hospital Capacity at Counties (Florida) Based on the Hypothetical Scenarios

In summary, the framework proposed for understanding and quantifying hospitalization rate can allow policy makers to a) evaluate the impact of COVID-19 virus transmission on hospitalization and ICU rates while controlling for demographics, health indicators and mobility trends so as to identify vulnerable counties that need to prioritized for vaccination; b) estimate the hospitalization and ICU rates at the county, state level to identify COVID-19 transmission trends under a host of future scenarios (c) Identify vulnerable locations with low projected hospital and ICU capacity (County/State/Region) in advance so that remedial measures can be adopted to provide additional hospital infrastructure and access to personnel and (d) Develop a comprehensive plan for assisting hospital systems across the country to address displaced demand due to COVD-19.

To be sure, the paper is not without limitations. The data on the hospitalization rate are continuously updated for few counties to correct for errors or omission. The models developed were based on the latest versions of the data at the time of manuscript preparation. The scenario analysis assumes future mobility to be similar across the scenarios. It is possible that mobility might also be affected as virus transmission rates change. The model is intended to serve as skeletal framework that can be readily updated with newer data on virus transmission and mobility patterns.

## Data Availability

All the data is available and the associated link is provided in the main manuscript and reference section.

## Contributors

NE conceptualized the study. TB and NE finalized the study design. TB conducted the literature review. TB collected the data. TB and NE analyzed and interpreted the model results. TB and NE prepared the figures. TB and NE drafted the main manuscript. Both authors reviewed the results and approved the final version of the manuscript.

## Acknowledgement

The authors would like to gratefully acknowledge SafeGraph COVID-19 Data Consortium, Department of Health and Human services, County Health Ranking and Road Maps, Centers for Disease Control System for providing access to the data at county level for United States.

## Footnotes

1 To illustrate the applicability of our model, we also evaluate the predictive performance of our proposed model in predicting all four response variable in context. The results are available upon request from the authors.

