## Supplemental Material for "A Comprehensive County Level Framework to Identify Factors Affecting Hospital Capacity and Predict Future Hospital Demand"

### *A1. Non-COVID Hospitalization Model*

COVID-19 Related Factors: As hypothesized, we find that an increase in weekly COVID transmission rate in the county results in a decrease in the non-COVID hospitalization rate. The result highlights how COVID-19 cases are affecting the general population's perception of hospital safety. It is also possible that hospitals are delaying hospitalization of other non-emergency patients to allow for requisite beds for COVID patients.

Demographics: As expected, counties with higher percentage of young individuals are less likely to experience higher non-COVID hospitalization rate. Similar to the COVID hospitalization trend, we find increased presence of minority population including Hispanic and African-American people in a county significantly increases hospitalization risk. Finally, our results show that women usually have a high hospitalization rate compared to men.

Health Indicators: Consistent with previous research, we also find that people suffering from pre-existing chronic diseases including cancer and HIV significantly increase the risk of being hospitalized.

Spatial Factors: With respect to spatial factors, we observe that mid-west region is more likely to have higher number of non-COVID hospitalization rates relative to other regions.

Temporal Factors: Similar to the COVID hospitalization model, we tested for the influence of temporal variables on the non-COVID hospitalization rate. We did not find any influence of the indicator variable from October 30$^{th}$ in the model. However, we did find the indicator variable from December 25$^{th}$ providing a mirror image of the results from COVID hospitalization rates. To elaborate, the variable reveals a negative coefficient indicating a reduced likelihood of the number of non-COVID patients across the country since 25$^{th}$ December. This variable directly reflects the influx of COVID patients reducing hospital capacity for non-COVID patients.

Correlation Factors: Similar to the COVID hospitalization rate, we also find the presence of common unobserved factors influencing county non-COVID hospitalization rate.

### *A2. ICU Usage Model (COVID and Non-COVID)*

Discussion about the ICU usage model will be provided upon request from the authors.

**Table A.1:** ICU Model Results

| | **COVID** | | **Non COVID** | |
|---|---|---|---|---|
| Parameter | Estimate | t-statistics | Estimate | t-statistics |
| Intercept | -11·038 | -8·612 | -20·854 | -10·978 |
| ***Covid-19 Related Factors*** | | | | |
| COVID case per 100 people, with 1 week lag | --** | -- | -0·056 | -3·044 |
| COVID case per 100 people, with 2 weeks lag | 0·659 | 13·045 | -0·054 | -2·859 |
| x Effect in Mid-West Region | -0·260 | -4·440 | -- | -- |
| x Effect in South Region | -0·324 | -5·762 | -- | -- |
| x Effect in North-east Region | | | -0·169 | -2·018 |
| % difference from 3 week moving average | 0·037 | 3·347 | -- | -- |
| x Effect in Mid-West Region | 0·022 | 1·732 | -- | -- |
| Weekly Covid-19 cases higher than the moving average (base is covid-19 cases same or lower) | 0·017 | 2·346 | -- | -- |
| ***Mobility Trends*** | | | | |
| Ln (Daily Average Exposure), 2 weeks lag | 0·085 | 5·430 | -- | -- |
| x Effect Since 2nd Wave started (October 30th ) | 0·036 | 10·382 | -- | -- |
| ***Demographics*** | | | | |
| Young people percentage | -- | -- | -0·043 | -4·555 |
| Hispanic people percentage | 0·015 | 9·344 | 0·013 | 5·028 |
| African American percentage | 0·013 | 7·538 | -- | -- |
| x Effect Since 2nd Wave started (October 30th ) | -0·002 | -2·319 | -- | -- |
| Female percentage | 0·129 | 10·639 | 0·206 | 12·038 |
| Income inequality ratio | 0·110 | 3·634 | -- | -- |
| ***Health Indicators*** | | | | |
| Ln (number of cardiovascular patients per 1000 Medicare beneficiaries) | 0·372 | 4·353 | 0·887 | 6·721 |
| Ln (HIV rate per 100K People) | -- | -- | 0·346 | 11·304 |
| Ln (cancer rate per 100K People) | 0·397 | 1·897 | 1·228 | 4·044 |
| ***Spatial Factors*** | | | | |
| Region (Base: South, Mid-west, Pacific) | | | | |
| West region | -- | -- | 0·443 | 4·599 |
| North East region | -- | -- | -- | -- |
| x Effect Since 2nd Wave started (October 30th ) | 0·064 | 1·777 | -- | -- |
| ***Temporal Factors*** | | | | |
| Effect Since 25th December | -- | -- | -0·021 | -2·095 |
| ***Correlations*** | | | | |
| $\sigma^2$ | 1·060 | 49·154 | 1·535 | 37·862 |
| $\rho$ | 0·931 | 468·920 | 0·982 | 325·480 |
| $\Phi$ | 0·848 | 260·942 | 0·881 | 267·780 |

** the variable is insignificant at 90% significance level.

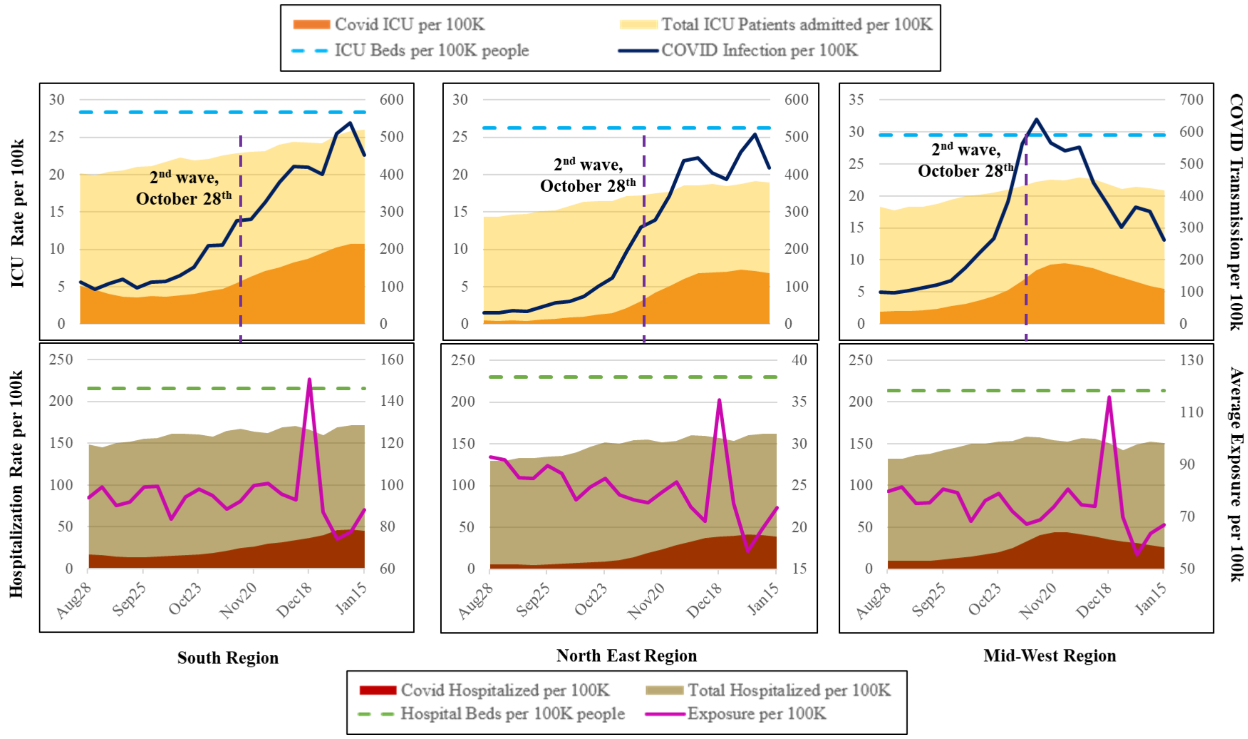


**Figure A.1:** A Representation of the Hospitalization Trend Across South, North-East and Mid-West region

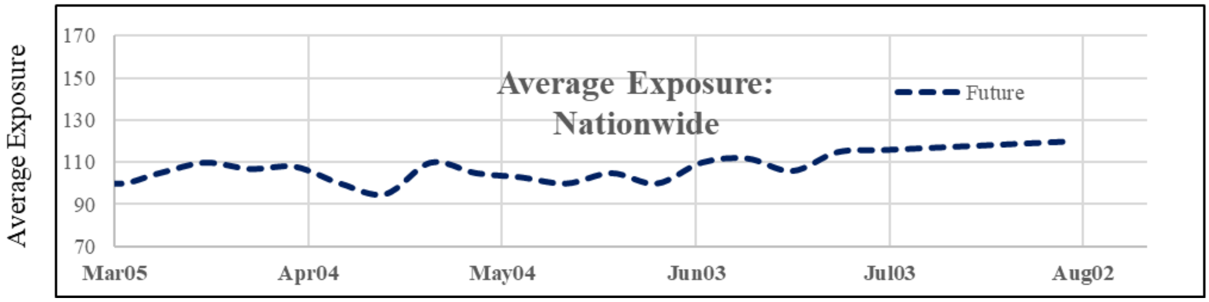


**Figure A.2:** Assumed Average Exposure Trend in Future

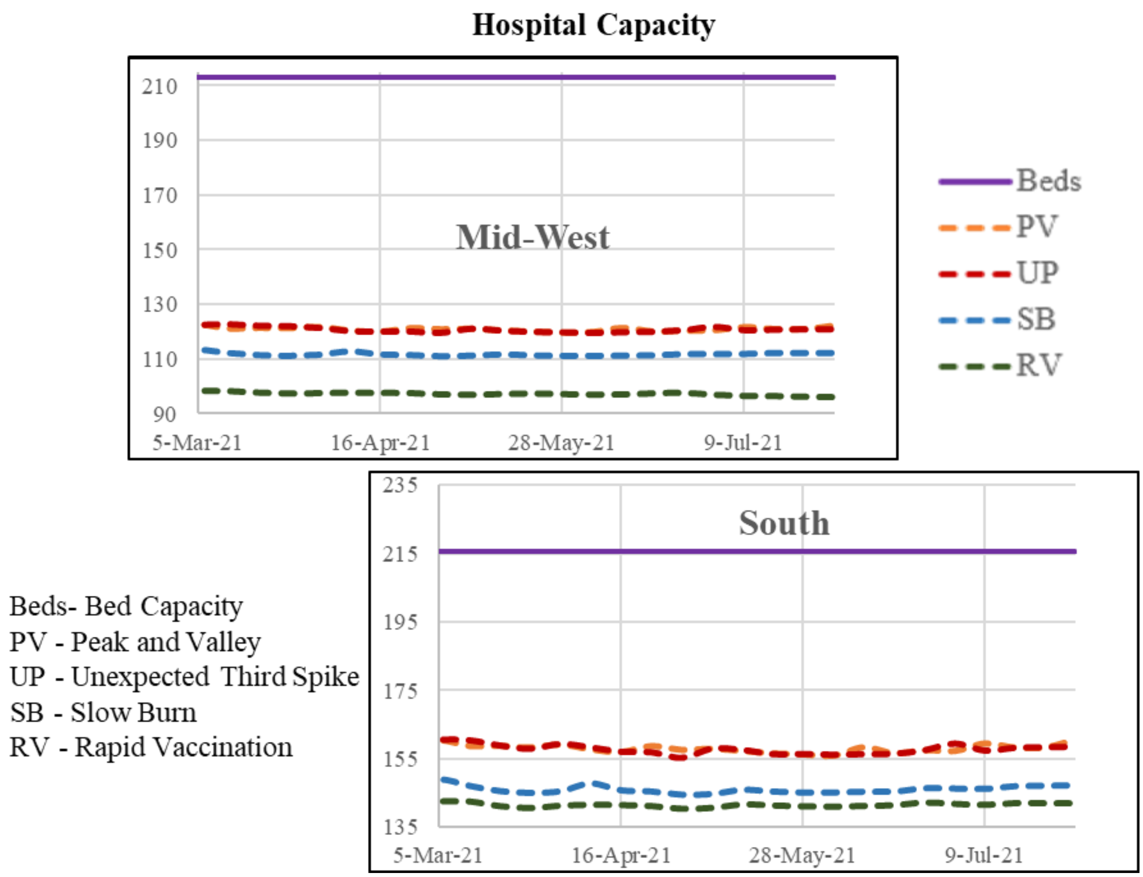


**Figure A.3:** Future Hospital Capacity Across Mod-West and South Regions Based on the Hypothetical Scenarios

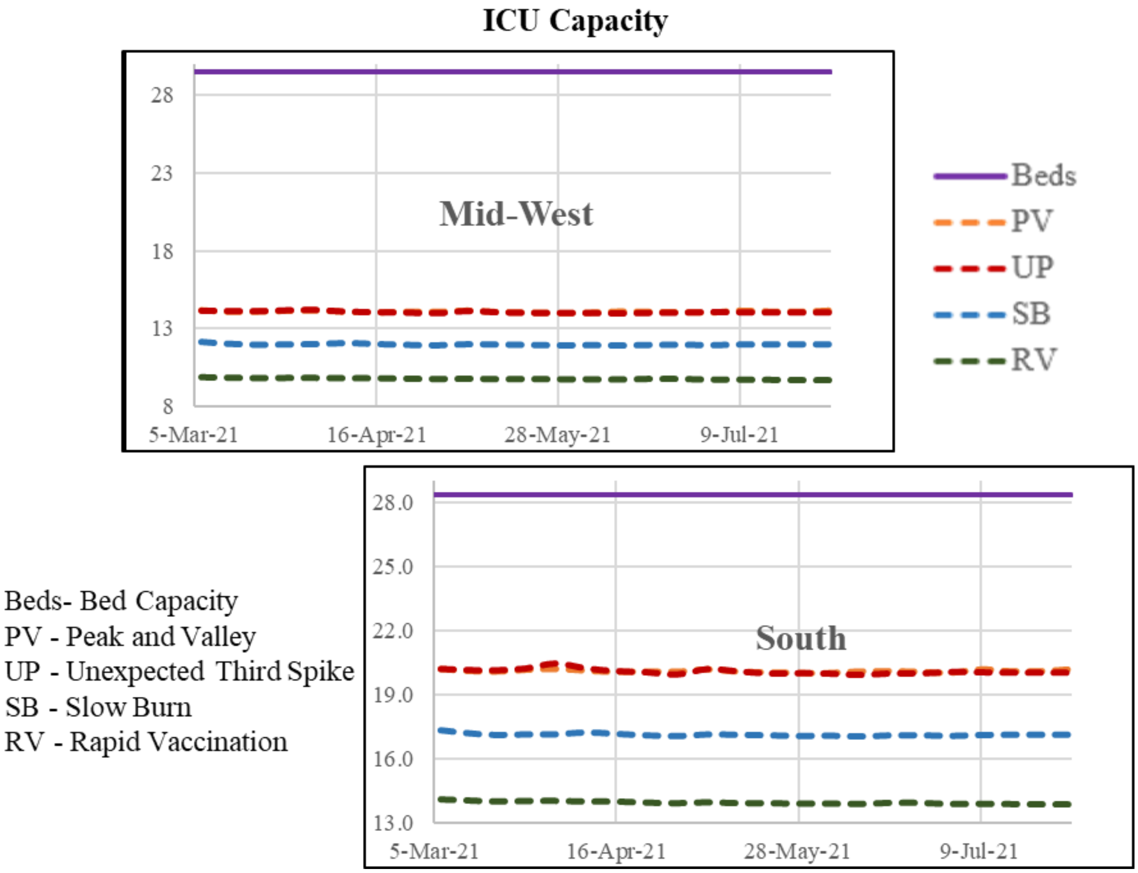


**Figure A.4:** Future ICU Capacity Across Mod-West and South Regions Based on the Hypothetical Scenarios

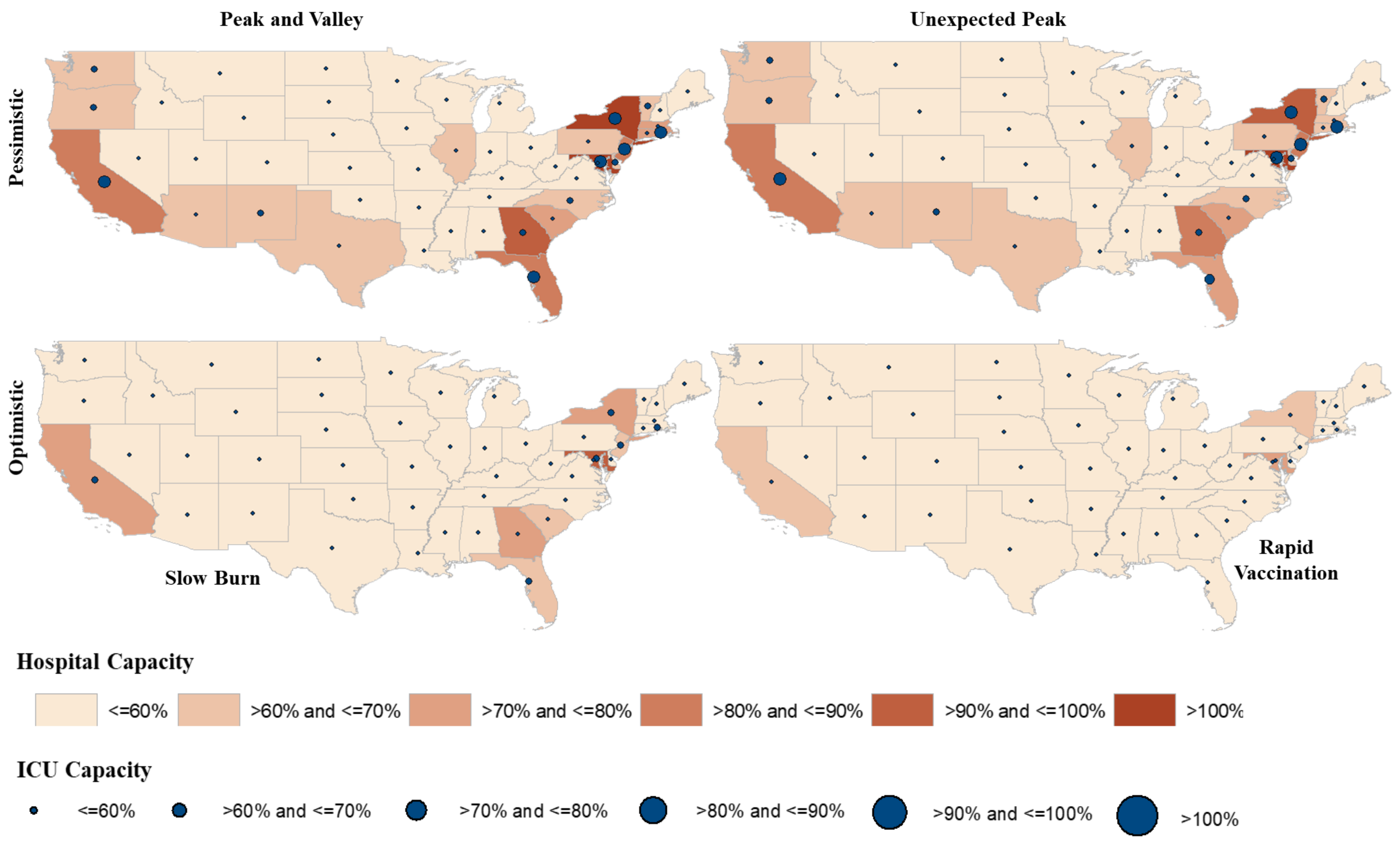


**Figure A.5:** Future Hospital Capacity at State level Based on the Hypothetical Scenarios
